## Additional File 1 for "Admixed and single-continental genome segments of the same ancestry have distinct linkage disequilibrium patterns"

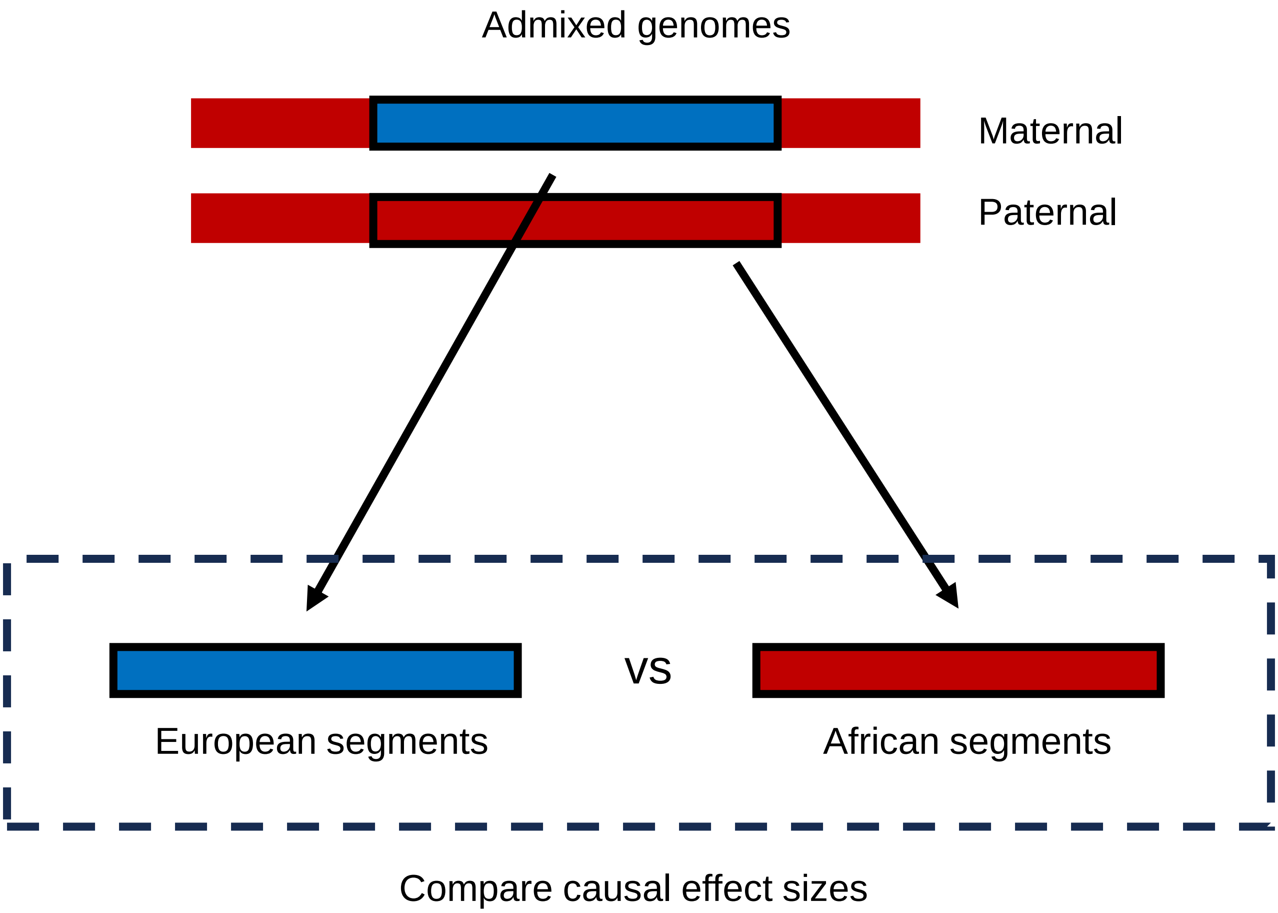


**Fig S1**. Overview of the analysis of Hou et al. (2023). African and European segments only from admixed genomes were compared to each other.

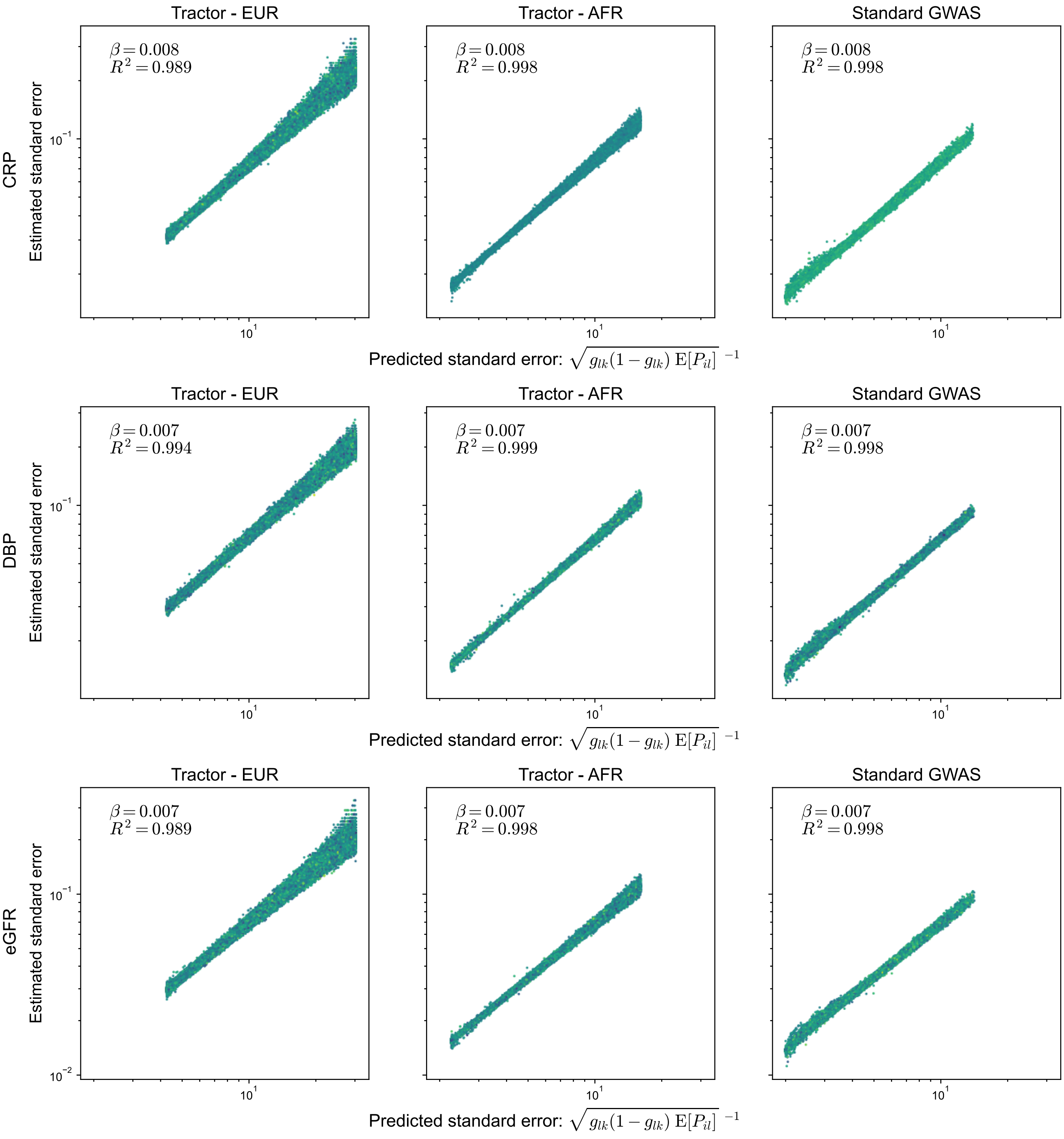


**Fig S2**. Estimated versus predicted standard error of Tractor and standard GWAS regression coefficients or three quantitative traits. The traits are on the left most of the figure.

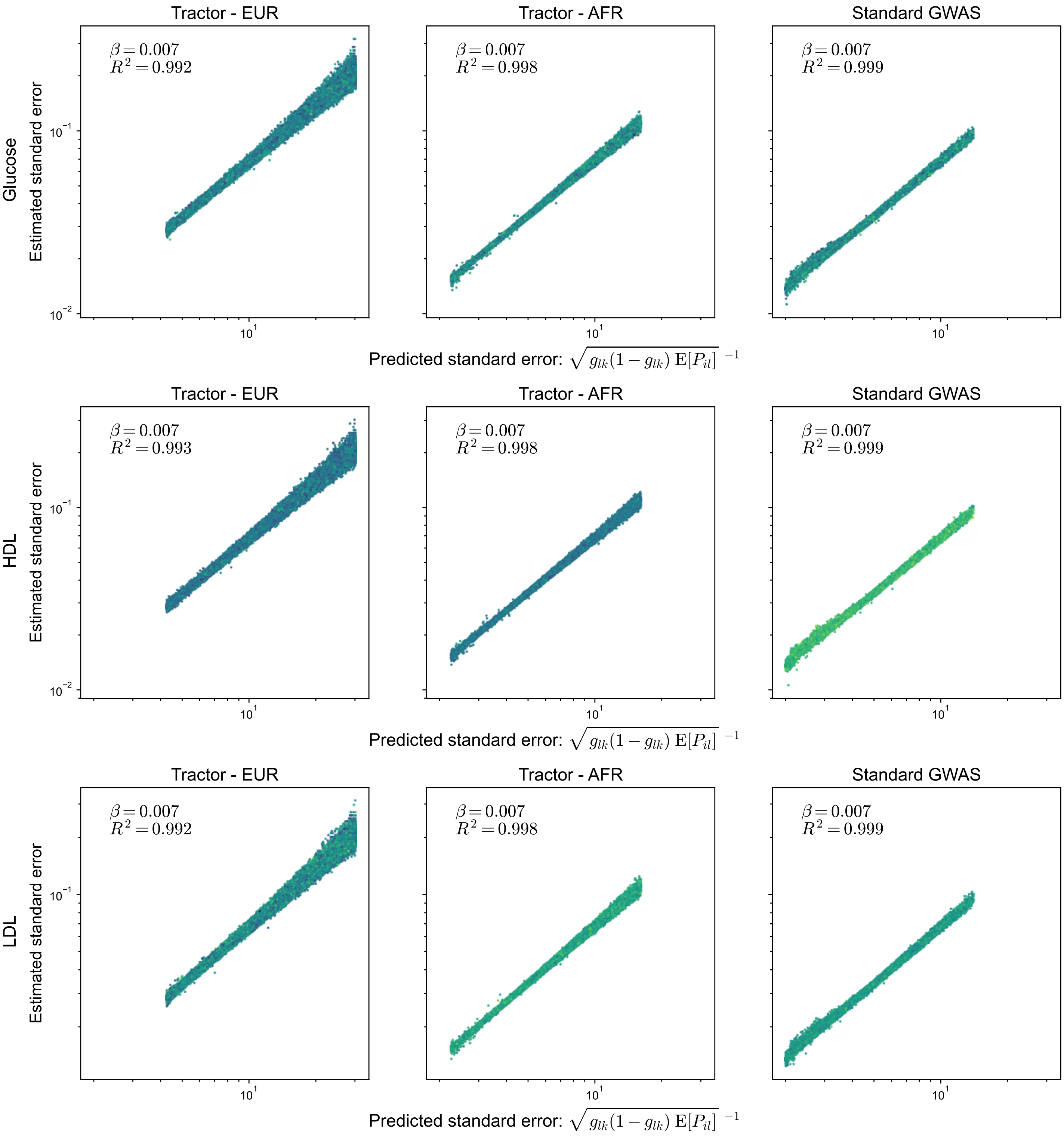


**Fig S3**. Estimated versus predicted standard error of Tractor and standard GWAS regression coefficients or three quantitative traits. The traits are on the left most of the figure.

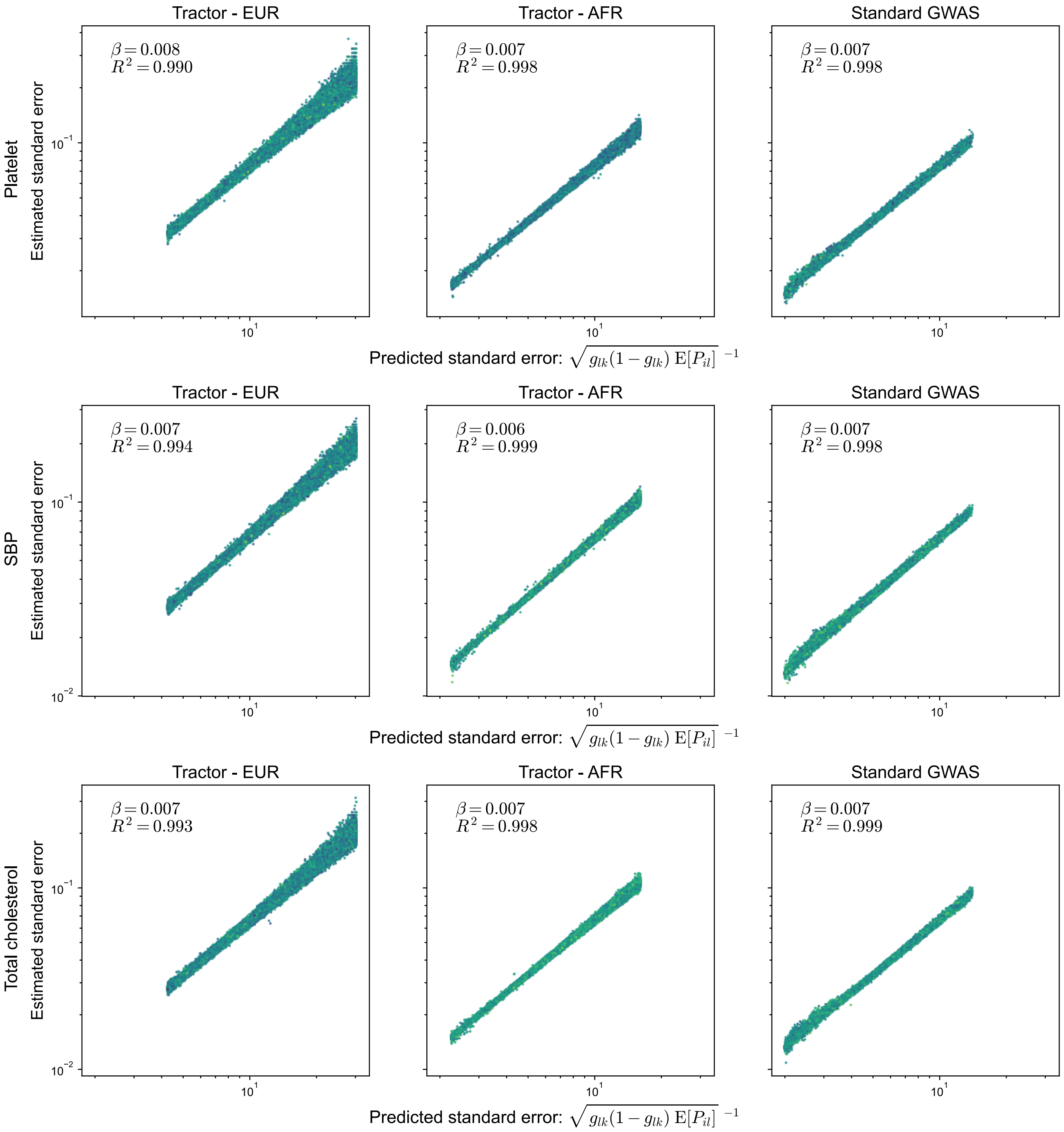


**Fig S4**. Estimated versus predicted standard error of Tractor and standard GWAS regression coefficients or three quantitative traits. The traits are on the left most of the figure.

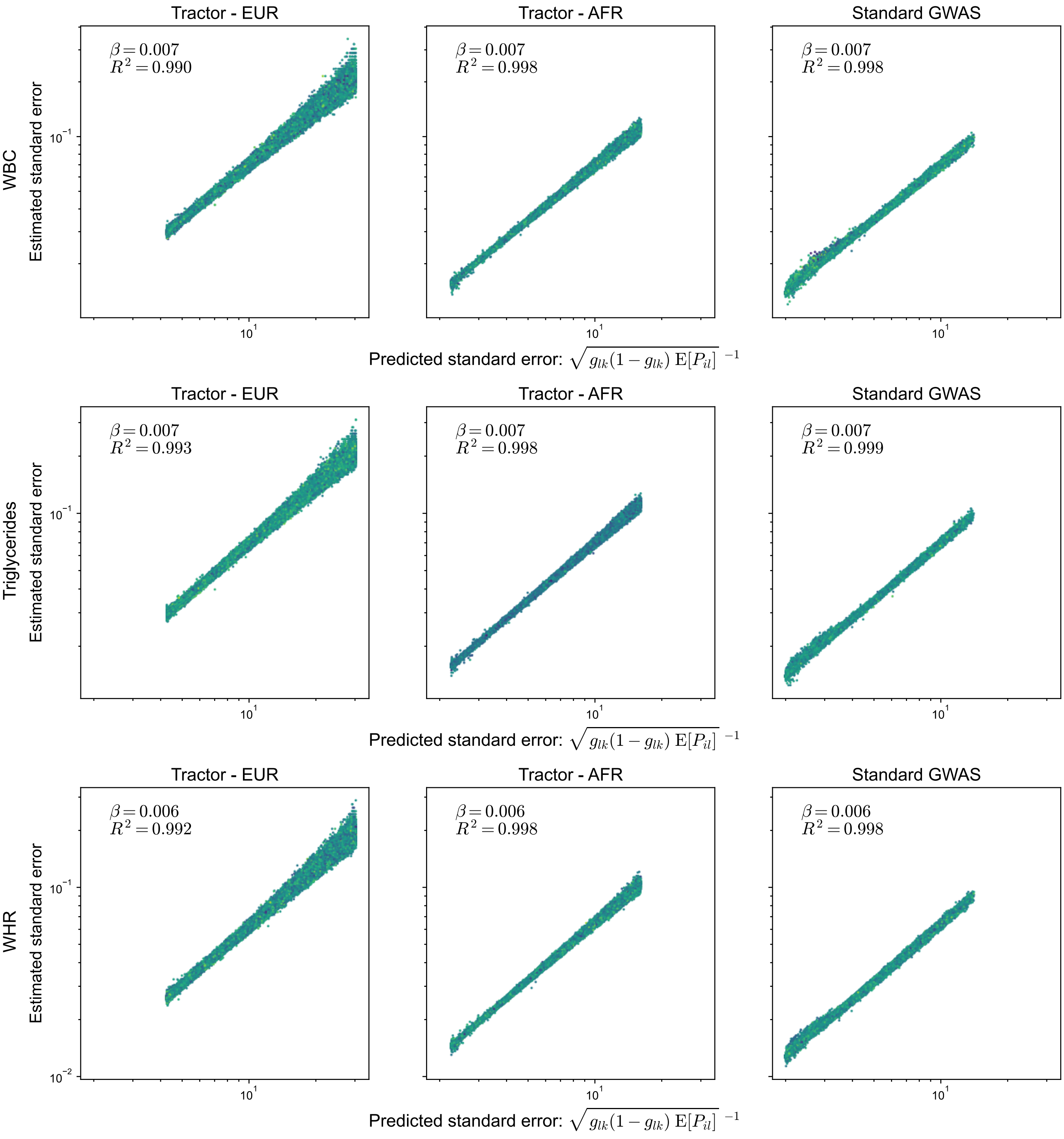


**Fig S5**. Estimated versus predicted standard error of Tractor and standard GWAS regression coefficients or three quantitative traits. The traits are on the left most of the figure.

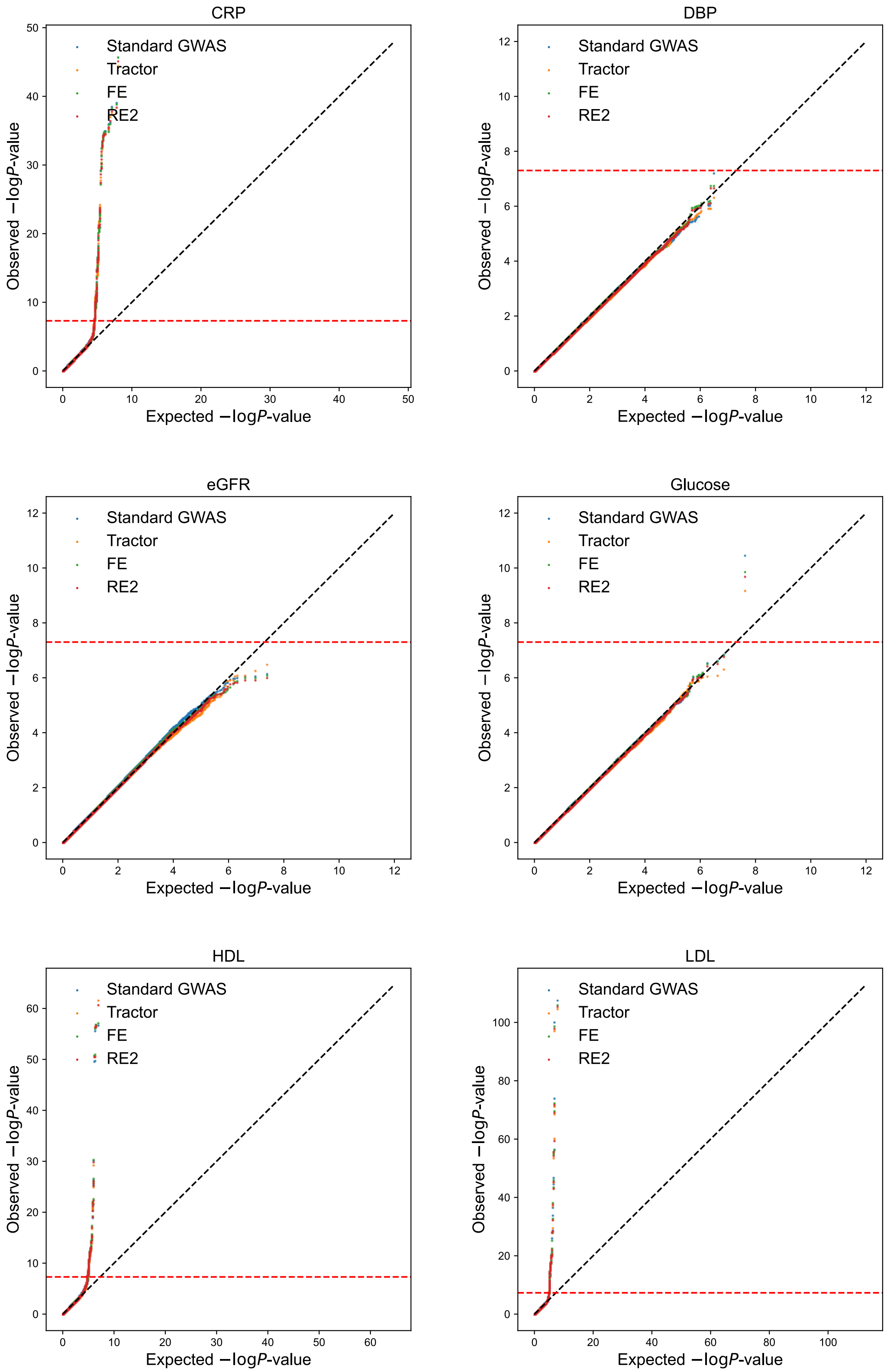


**Fig S6**. QQ-plots of 6 quantitative traits: C-reactive protein, diastolic blood pressure, estimated GFR, glucose, HDL, and LDL.

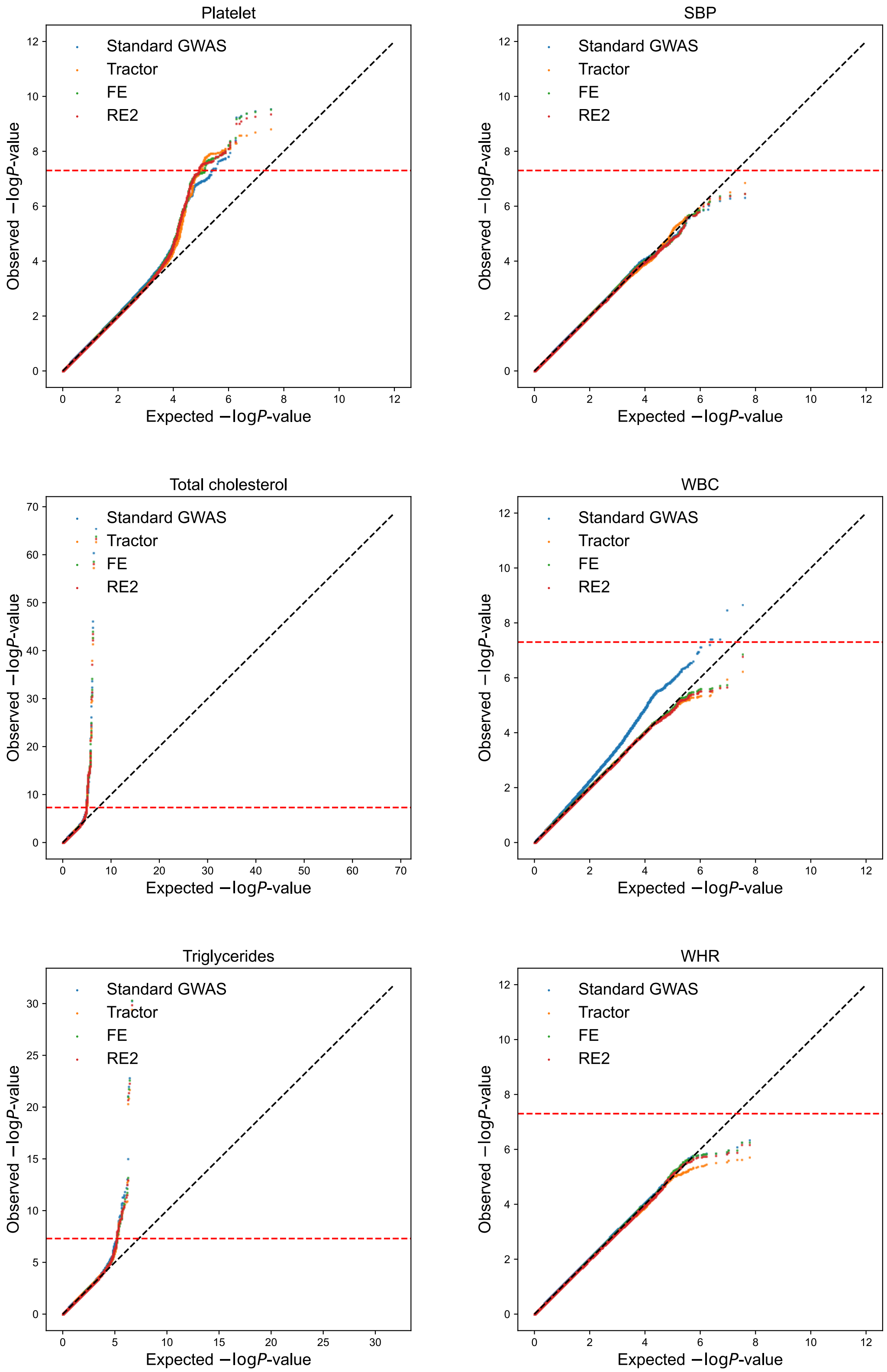


**Fig S7**. QQ-plots of 6 quantitative traits: Platelet count, systolic blood pressure, total cholesterol, white blood cell count, triglycerides, and waist-to-hip ratio.

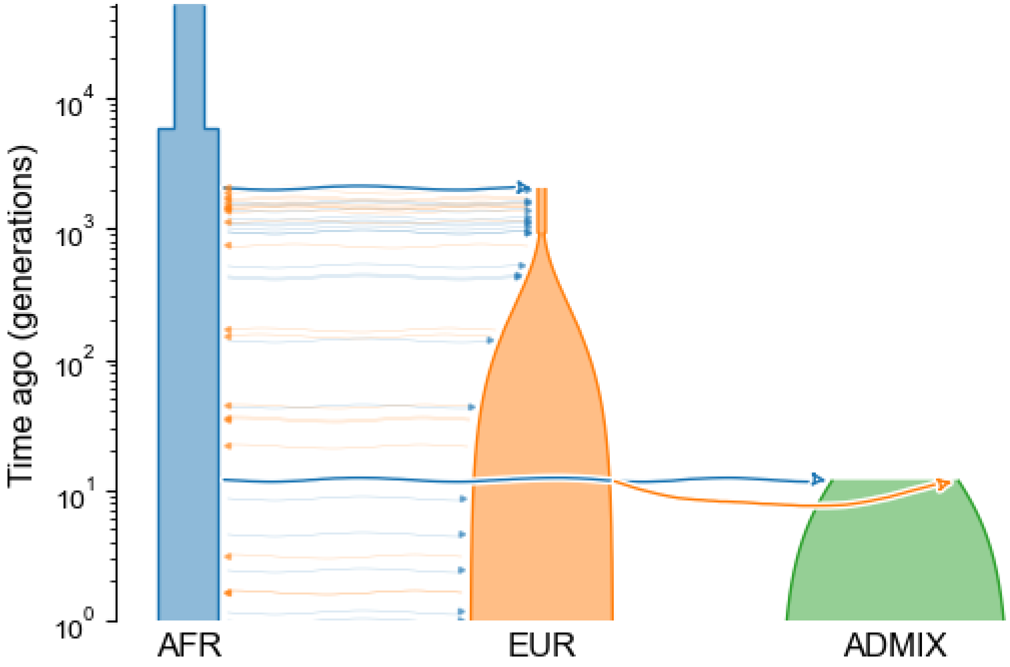


**Fig S8**. The demographic model of African-American admixture adopted from Browning et al. (2018). The demes yaml file was retrieved from stdpopsim catalog.
Link: https://popsim-consortium.github.io/stdpopsim-docs/stable/catalog.html
