## Additional File 2 for "Admixed and single-continental genome segments of the same ancestry have distinct linkage disequilibrium patterns"

### Additional File 2: Supplementary Note

#### Proof of Equation 11

**Equation 11** follows from the following statements (**Equation 26** and **Equation 27**). Note that **Equation 11** assumes the case of two ancestries, i.e., $l = 1, 2$).

$\mathbb{E}\left[\widetilde{\mathbf{M}}_{ik}^T \widetilde{\mathbf{M}}_{ik}\right]$ can be calculated as follows:

$$\begin{aligned}
\mathbb{E}\left[\widetilde{\mathbf{M}}_{ik}^T \widetilde{\mathbf{M}}_{ik}\right] &= \mathrm{Var}(\mathbf{M}_{ik} - \mathbb{E}[\mathbf{M}_{ik} \mid \mathbf{L}_{ik}]) \\
&= \mathbb{E}[\mathrm{Var}(\mathbf{M}_{ik} \mid \mathbf{L}_{ik})] \\
&= 2\mathbb{E}[\mathrm{Var}(\mathbf{M}_{ik}^{\mathbf{h}} \mid \mathbf{L}_{ik})] \quad \because M_{ik}^{\mathbf{m}} \perp\!\!\!\perp M_{ik}^{\mathbf{p}} \text{ (HWE)} \\
&= \begin{bmatrix} 2g_{1k}(1-g_{1k})\mathbb{E}[P_{i1}] & \cdots & 0 \\ \vdots & \ddots & \vdots \\ 0 & \cdots & 2g_{n_L k}(1-g_{n_L k})\mathbb{E}[P_{in_L}] \end{bmatrix}
\end{aligned} \tag{26}$$

Note that only diagonal elements are non-zero.

The remaining term, $\mathbb{E}\left[\widetilde{M}_{ik}^2\right]$, is calculated as follows.

$$\begin{aligned}
\mathbb{E}\left[\widetilde{M}_{ik}^2\right] &= \mathbb{E}[\mathrm{Var}(M_{ik} \mid \mathbf{P}_i)] \\
&= 2\mathbb{E}[\mathrm{Var}(M_{ik}^{\mathbf{h}} \mid \mathbf{P}_i)] \quad \because M_{ik}^{\mathbf{m}} \perp\!\!\!\perp M_{ik}^{\mathbf{p}} \text{ (HWE)}
\end{aligned} \tag{27}$$

**Equation 28** at the next subsection finishes the proof. Since we assume the case of two ancestries, $P_{i1} + P_{i2} = 1$, hence $P_{i1}(1 - P_{i1}) = P_{i2}(1 - P_{i2}) = P_{i1}P_{i2}$, thereby yielding **Equation 11**.

#### Proof of Equation 18

**Equation 18** follows from the following statements (**Equation 28** and **Equation 29**). The law of total variance and the law of total covariance are applied, respectively.

The variance of $M_{ik}$ conditional to global ancestry, $\mathrm{Var}(M_{ik} \mid \mathbf{P}_i)$, is calculated as follows:

$$
\begin{aligned}
\mathrm{Var}(M_{ik} \mid \mathbf{P}_i) &= 2\mathrm{Var}(M^{\mathbf{h}}_{ik} \mid \mathbf{P}_i) \\
&= 2\Big[\mathrm{Var}(\mathbb{E}[M^{\mathbf{h}}_{ik} \mid \mathbf{L}^{\mathbf{h}}_{ik}, \mathbf{P}_i] \mid \mathbf{P}_i) + \mathbb{E}[\mathrm{Var}(M^{\mathbf{h}}_{ik} \mid \mathbf{L}^{\mathbf{h}}_{ik}, \mathbf{P}_i) \mid \mathbf{P}_i]\Big] \\
&= 2\Big[\mathrm{Var}(\mathbb{E}[M^{\mathbf{h}}_{ik} \mid \mathbf{L}^{\mathbf{h}}_{ik}] \mid \mathbf{P}_i) + \mathbb{E}[\mathrm{Var}(M^{\mathbf{h}}_{ik} \mid \mathbf{L}^{\mathbf{h}}_{ik}) \mid \mathbf{P}_i]\Big] \\
&= 2\left[\mathrm{Var}\Big(\sum_{l=1}^{n_L} g_{lk} L^{\mathbf{h}}_{ikl} \mid \mathbf{P}_i\Big) + \mathbb{E}\Big[\sum_{l=1}^{n_L} g_{lk}(1-g_{lk}) L^{\mathbf{h}}_{ikl} \mid \mathbf{P}_i\Big]\right] \\
&= 2\left[\sum_{l,l'} g_{lk} g_{l'k} \mathrm{Cov}(L^{\mathbf{h}}_{ikl}, L^{\mathbf{h}}_{ikl'} \mid \mathbf{P}_i) + \sum_{l=1}^{n_L} g_{lk}(1-g_{lk}) \mathbb{E}[L^{\mathbf{h}}_{ikl} \mid \mathbf{P}_i]\right] \\
&= 2\left[\sum_{l=1}^{n_L} g^2_{lk} P_{il}(1-P_{il}) + \sum_{l \neq l'} g_{lk} g_{l'k}(-P_{il}P_{il'}) + \sum_{l=1}^{n_L} g_{lk}(1-g_{lk}) P_{il}\right] \\
&= 2\sum_{l=1}^{n_L} g_{lk}(1-g_{lk}) P_{il} + 2\sum_{l=1}^{n_L} g^2_{lk} P_{il}(1-P_{il}) - 2\sum_{l \neq l'} g_{lk} g_{l'k} P_{il} P_{il'}
\end{aligned} \tag{28}
$$

The covariance of $M_{ik}$ and $C_{ij}$ conditional to global ancestry, $\mathrm{Cov}(M_{ik}, C_{ij} \mid \mathbf{P}_i)$, is calculated as follows:

$$
\begin{aligned}
\mathrm{Cov}(M_{ik}, C_{ij} \mid \mathbf{P}_i) &= 2\mathrm{Cov}(M^{\mathbf{h}}_{ik}, C^{\mathbf{h}}_{ij} \mid \mathbf{P}_i) \\
&= 2\mathbb{E}[\mathrm{Cov}(M^{\mathbf{h}}_{ik}, C^{\mathbf{h}}_{ij} \mid \mathbf{P}_i, \mathbf{L}^{\mathbf{h}}_{ik}, \mathbf{L}^{\mathbf{h}}_{ij}) \mid \mathbf{P}_i] \\
&+ 2\mathrm{Cov}(\mathbb{E}[M^{\mathbf{h}}_{ik} \mid \mathbf{P}_i, \mathbf{L}^{\mathbf{h}}_{ik}, \mathbf{L}^{\mathbf{h}}_{ij}], \mathbb{E}[C^{\mathbf{h}}_{ij} \mid \mathbf{P}_i, \mathbf{L}^{\mathbf{h}}_{ik}, \mathbf{L}^{\mathbf{h}}_{ij}] \mid \mathbf{P}_i) \\
&= 2\mathbb{E}[\mathrm{Cov}(M^{\mathbf{h}}_{ik}, C^{\mathbf{h}}_{ij} \mid \mathbf{L}^{\mathbf{h}}_{ik}, \mathbf{L}^{\mathbf{h}}_{ij}) \mid \mathbf{P}_i] \\
&+ 2\mathrm{Cov}(\mathbb{E}[M^{\mathbf{h}}_{ik} \mid \mathbf{L}^{\mathbf{h}}_{ik}], \mathbb{E}[C^{\mathbf{h}}_{ij} \mid \mathbf{L}^{\mathbf{h}}_{ij}] \mid \mathbf{P}_i)
\end{aligned} \tag{29}
$$

where the first term $2\mathbb{E}[\mathrm{Cov}(M^{\mathbf{h}}_{ik}, C^{\mathbf{h}}_{ij} \mid \mathbf{L}^{\mathbf{h}}_{ik}, \mathbf{L}^{\mathbf{h}}_{ij}) \mid \mathbf{P}_i]$ is further calculated as follows, distinguishing between cases where the causal and marker variants share identical ancestry:

$$
2\mathbb{E}[\mathrm{Cov}(M^{\mathbf{h}}_{ik}, C^{\mathbf{h}}_{ij} \mid \mathbf{L}^{\mathbf{h}}_{ik}, \mathbf{L}^{\mathbf{h}}_{ij}) \mid \mathbf{P}_i] = \begin{cases} 2\mathbb{E}\left[\sum_{l=1}^{n_L} D_{ljk} L^{\mathbf{h}}_{ikl} \mid \mathbf{P}_i\right] & : \quad \mathbf{L}^{\mathbf{h}}_{ik} = \mathbf{L}^{\mathbf{h}}_{ij} \\ 0 & : \quad \mathbf{L}^{\mathbf{h}}_{ik} \neq \mathbf{L}^{\mathbf{h}}_{ij} \end{cases} \tag{30}
$$

$$
2\mathbb{E}\left[\sum_{l=1}^{n_L} D_{ljk} L^{\mathbf{h}}_{ikl} \mid \mathbf{P}_i\right] = 2\sum_{l=1}^{n_L} D_{ljk} \mathbb{E}\left[L^{\mathbf{h}}_{ikl} \mid \mathbf{P}_i\right] = 2\sum_{l=1}^{n_L} D_{ljk} P_{il} \tag{31}
$$

The second term, $2\text{Cov}(\mathbb{E}[M^{\mathbf{h}}_{ik} \mid \mathbf{L}^{\mathbf{h}}_{ik}], \mathbb{E}[C^{\mathbf{h}}_{ij} \mid \mathbf{L}^{\mathbf{h}}_{ij}] \mid \mathbf{P}_i)$ is calculated as follows:

$$\begin{aligned}
2\text{Cov}(\mathbb{E}[M^{\mathbf{h}}_{ik} \mid \mathbf{L}^{\mathbf{h}}_{ik}], \mathbb{E}[C^{\mathbf{h}}_{ij} \mid \mathbf{L}^{\mathbf{h}}_{ij}] \mid \mathbf{P}_i) &= 2\text{Cov}\left(\sum_{l=1}^{n_L} g_{lk} L^{\mathbf{h}}_{ikl}, \sum_{l=1}^{n_L} f_{lj} L^{\mathbf{h}}_{ijl} \mid \mathbf{P}_i\right) \\
&= 2\sum_{l,l'} g_{lk} f_{l'j} \text{Cov}(L^{\mathbf{h}}_{ikl}, L^{\mathbf{h}}_{ijl'} \mid \mathbf{P}_i) \\
&= 2\sum_{l=1}^{n_L} g_{lk} f_{lj} P_{il}(1 - P_{il}) - 2\sum_{l \neq l'} g_{lk} f_{l'k} P_{il} P_{il'}
\end{aligned} \tag{32}$$

Combining the above equations completes the proof, thereby yielding **Equation 18**.

#### Proof of Equation 19

**Equation 19** follows from the following statements:

$$\begin{aligned}
\mathbb{E}[Y_i \mid \mathbf{M}_{ik}, \mathbf{L}_{ik}, \mathbf{P}_i] &= \mathbb{E}\left[\alpha_0 + \sum_{j=1}^{n_J} C_{ij}\alpha_j + \varepsilon_i \mid \mathbf{M}_{ik}, \mathbf{L}_{ik}, \mathbf{P}_i\right] \\
&= \alpha_0 + \sum_{j=1}^{n_J} \mathbb{E}[C_{ij} \mid \mathbf{M}_{ik}, \mathbf{L}_{ik}, \mathbf{P}_i]\alpha_j \quad \because M_{ik} \perp\!\!\!\perp \varepsilon_i \mid \mathbf{P}_i \\
&= \alpha_0 + \sum_{j \in [k]_l} \mathbb{E}[C_{ij} \mid \mathbf{M}_{ik}, \mathbf{L}_{ik}, \mathbf{P}_i]\alpha_j + \sum_{j \notin [k]_l} \mathbb{E}[C_{ij} \mid \mathbf{M}_{ik}, \mathbf{L}_{ik}, \mathbf{P}_i]\alpha_j \\
&= \alpha_0 + \sum_{j \in [k]_l} \mathbb{E}[C_{ij} \mid \mathbf{M}_{ik}, \mathbf{L}_{ik}]\alpha_j + \sum_{j \notin [k]_l} \mathbb{E}[C_{ij} \mid \mathbf{P}_i]\alpha_j
\end{aligned} \tag{33}$$

where

$$\begin{aligned}
\mathbb{E}[C_{ij} \mid \mathbf{M}_{ik}, \mathbf{L}_{ik}] &= 2\mathbb{E}[C^{\mathbf{h}}_{ij} \mid \mathbf{M}_{ik}, \mathbf{L}_{ik}] \\
&= 2\mathbb{P}(C^{\mathbf{h}}_{ij} = 1 \mid \mathbf{M}_{ik}, \mathbf{L}_{ik}) \\
&= 2 \cdot \frac{\mathbb{P}(C^{\mathbf{h}}_{ij} = 1, \mathbf{M}_{ik} \mid \mathbf{L}_{ik})}{\mathbb{P}(\mathbf{M}_{ik} \mid \mathbf{L}_{ik})}
\end{aligned} \tag{34}$$

Given that $\sum_{l=1}^{n_L} L_{ikl} = 2$, only two cases arise: (1) $L_{ikl} = 2$ for some $l$ and $L_{ikl'} = 0$ for all $l' \neq l$, or (2) $L_{ikl} = L_{ikl'} = 1$ for some $l, l'$ and 0 otherwise. Therefore, it suffices to prove for the two following *simplified* cases: (1) $n_L = 1$ where $L_{ik1} = 2$, and (2) $n_L = 2$ where $(L_{ik1}, L_{ik2}) = (1, 1)$.

**Case 1:** $n_L = 1$, where $L_{ik1} = 2$. The possible values for $M_{ik1}$ are 0, 1, and 2.

$$\begin{aligned}
2 \cdot \frac{\mathbb{P}(C_{ij}^{\mathbf{h}} = 1, \mathbf{M}_{ik} \mid \mathbf{L}_{ik})}{\mathbb{P}(\mathbf{M}_{ik} \mid \mathbf{L}_{ik})} &= 2 \cdot \frac{\mathbb{P}(C_{ij}^{\mathbf{h}} = 1, M_{ik1} \mid \mathbf{L}_{ik})}{\mathbb{P}(M_{ik1} \mid \mathbf{L}_{ik})} \\
&= \begin{cases} 2 \cdot \frac{f_{1j} - h_{1jk}}{1 - g_{1k}} & : M_{ik1} = 0 \\ \frac{f_{1j} - h_{1jk}}{1 - g_{1k}} + \frac{h_{1jk}}{g_{1k}} & : M_{ik1} = 1 \\ 2 \cdot \frac{h_{1jk}}{g_{1k}} & : M_{ik1} = 2 \end{cases} \\
&= \frac{D_{1jk}}{g_{1k}(1 - g_{1k})} M_{ik1} + 2 \cdot \frac{f_{1j} - h_{1jk}}{1 - g_{1k}}
\end{aligned} \tag{35}$$

Note that $D_{1jk} = h_{1jk} - f_{1j} g_{1k}$.

A detailed proof for **Equation 35** is provided in **Equation 36** and **Equation 37**. Note that $\mathbb{P}(C_{ij}^{\mathbf{h}} = 1, M_{ik1} \mid \mathbf{L}_{ik}) = \mathbb{P}(C_{ij}^{\mathbf{h}} = 1, M_{ik1}^{\mathbf{h}} + M_{ik1}^{\mathbf{h}'} \mid \mathbf{L}_{ik})$, where $\mathbf{h} \neq \mathbf{h}'$.

$$\mathbb{P}(M_{ik1} = r \mid \mathbf{L}_{ik}) = \binom{2}{r} g_{1k}^{r} (1 - g_{1k})^{2-r} \quad (r = 0, 1, 2) \tag{36}$$

$$\begin{aligned}
\mathbb{P}(C_{ij}^{\mathbf{h}} = 1, M_{ik1} = 0 \mid \mathbf{L}_{ik}) &= \mathbb{P}(C_{ij}^{\mathbf{h}} = 1, M_{ik1}^{\mathbf{h}} = 0, M_{ik1}^{\mathbf{h}'} = 0 \mid \mathbf{L}_{ik}) \\
&= \mathbb{P}(C_{ij}^{\mathbf{h}} = 1, M_{ik1}^{\mathbf{h}} = 0 \mid \mathbf{L}_{ik}) \cdot \mathbb{P}(M_{ik1}^{\mathbf{h}'} = 0 \mid \mathbf{L}_{ik}) \quad \because \text{HWE} \\
&= (f_{1j} - h_{1jk})(1 - g_{1k}) \\
\mathbb{P}(C_{ij}^{\mathbf{h}} = 1, M_{ik1} = 1 \mid \mathbf{L}_{ik}) &= \mathbb{P}(C_{ij}^{\mathbf{h}} = 1, M_{ik1}^{\mathbf{h}} = 0, M_{ik1}^{\mathbf{h}'} = 1 \mid \mathbf{L}_{ik}) \\
&+ \mathbb{P}(C_{ij}^{\mathbf{h}} = 1, M_{ik1}^{\mathbf{h}} = 1, M_{ik1}^{\mathbf{h}'} = 0 \mid \mathbf{L}_{ik}) \\
&= \mathbb{P}(C_{ij}^{\mathbf{h}} = 1, M_{ik1}^{\mathbf{h}} = 0 \mid \mathbf{L}_{ik}) \cdot \mathbb{P}(M_{ik1}^{\mathbf{h}'} = 1 \mid \mathbf{L}_{ik}) \\
&+ \mathbb{P}(C_{ij}^{\mathbf{h}} = 1, M_{ik1}^{\mathbf{h}} = 1 \mid \mathbf{L}_{ik}) \cdot \mathbb{P}(M_{ik1}^{\mathbf{h}'} = 0 \mid \mathbf{L}_{ik}) \quad \because \text{HWE} \\
&= (f_{1j} - h_{1jk}) g_{1k} + h_{1jk}(1 - g_{1k}) \\
\mathbb{P}(C_{ij}^{\mathbf{h}} = 1, M_{ik1} = 2 \mid \mathbf{L}_{ik}) &= \mathbb{P}(C_{ij}^{\mathbf{h}} = 1, M_{ik1}^{\mathbf{h}} = 1, M_{ik1}^{\mathbf{h}'} = 1 \mid \mathbf{L}_{ik}) \\
&= \mathbb{P}(C_{ij}^{\mathbf{h}} = 1, M_{ik1}^{\mathbf{h}} = 1 \mid \mathbf{L}_{ik}) \cdot \mathbb{P}(M_{ik1}^{\mathbf{h}'} = 1 \mid \mathbf{L}_{ik}) \quad \because \text{HWE} \\
&= h_{1jk} \cdot g_{1k}
\end{aligned} \tag{37}$$

Dividing **Equation 37** by **Equation 36** yields **Equation 35**.

**Case 2:** $n_L = 2$ where $(L_{ik1}, L_{ik2}) = (1,1)$. The possible combinations for $M_{ik1}$ are (0,0), (1,0), (0,1), and (1,1).

$$
\begin{aligned}
2 \cdot \frac{\mathbb{P}(C_{ij}^{\mathbf{h}} = 1, \mathbf{M}_{ik} \mid \mathbf{L}_{ik})}{\mathbb{P}(\mathbf{M}_{ik} \mid \mathbf{L}_{ik})} &= 2 \cdot \frac{\mathbb{P}(C_{ij}^{\mathbf{h}} = 1, M_{ik1}, M_{ik2} \mid \mathbf{L}_{ik})}{\mathbb{P}(M_{ik1}, M_{ik2} \mid \mathbf{L}_{ik})} \\
&= \begin{cases} \frac{f_{1j}-h_{1jk}}{1-g_{1k}} + \frac{f_{2j}-h_{2jk}}{1-g_{2k}} & : (M_{ik1}, M_{ik2}) = (0,0) \\ \frac{h_{1jk}}{g_{1k}} + \frac{f_{2j}-h_{2jk}}{1-g_{2k}} & : (M_{ik1}, M_{ik2}) = (1,0) \\ \frac{f_{1j}-h_{1jk}}{1-g_{1k}} + \frac{h_{2jk}}{g_{2k}} & : (M_{ik1}, M_{ik2}) = (0,1) \\ \frac{h_{1jk}}{g_{1k}} + \frac{h_{2jk}}{g_{2k}} & : (M_{ik1}, M_{ik2}) = (1,1) \end{cases} \\
&= \sum_{l=1,2} \frac{D_{ljk}}{g_{lk}(1-g_{lk})} M_{ikl} + \sum_{l=1,2} \frac{f_{lj} - h_{ljk}}{1 - g_{lk}} L_{ikl}
\end{aligned} \tag{38}
$$

A detailed proof is provided in **Equation 39** and **Equation 40**. Note that $s = 0, 1$ and $t = 0, 1$.

$$
\mathbb{P}(M_{ik1} = s, M_{ik2} = t \mid \mathbf{L}_{ik}) = 2(g_{1k})^s (1 - g_{1k})^{1-s} (g_{2k})^t (1 - g_{2k})^{1-t} \tag{39}
$$

$$
\begin{aligned}
\mathbb{P}(C_{ij}^{\mathbf{h}} = 1, M_{ik1} = s, M_{ik2} = t \mid \mathbf{L}_{ik}) &= \mathbb{P}(C_{ij1}^{\mathbf{h}} = 1, M_{ik1}^{\mathbf{h}} = s, M_{ik2}^{\mathbf{h}'} = t \mid \mathbf{L}_{ik}) \\
&+ \mathbb{P}(C_{ij2}^{\mathbf{h}} = 1, M_{ik2}^{\mathbf{h}} = t, M_{ik1}^{\mathbf{h}'} = s \mid \mathbf{L}_{ik}) \\
&= \mathbb{P}(C_{ij1}^{\mathbf{h}} = 1, M_{ik1}^{\mathbf{h}} = s \mid \mathbf{L}_{ik}) \cdot \mathbb{P}(M_{ik2}^{\mathbf{h}'} = t \mid \mathbf{L}_{ik}) \\
&+ \mathbb{P}(C_{ij2}^{\mathbf{h}} = 1, M_{ik2}^{\mathbf{h}} = t \mid \mathbf{L}_{ik}) \cdot \mathbb{P}(M_{ik1}^{\mathbf{h}'} = s \mid \mathbf{L}_{ik}) \quad \because \text{HWE} \\
&= (h_{1jk})^s (f_{1j} - h_{1jk})^{1-s} (g_{2k})^t (1 - g_{2k})^{1-t} \\
&+ (h_{2jk})^t (f_{2j} - h_{2jk})^{1-t} (g_{1k})^s (1 - g_{1k})^{1-s}
\end{aligned} \tag{40}
$$

Dividing **Equation 40** by **Equation 39** yields **Equation 38**. Additionally, both **Equation 37** and **Equation 40** can be generalized to the $n_L \geq 2$ case, assuming $\sum_{l=1,2} L_{ikl} = 2$. Finally, substituting $L_{ik1} = 2 - L_{ik2}$ yields the generalized result.

$$
\begin{aligned}
2 \cdot \frac{\mathbb{P}(C_{ij}^{\mathbf{h}} = 1, \mathbf{M}_{ik} \mid \mathbf{L}_{ik})}{\mathbb{P}(\mathbf{M}_{ik} \mid \mathbf{L}_{ik})} &= \sum_{l=1}^{n_L} \frac{D_{ljk}}{g_{lk}(1-g_{lk})} M_{ikl} \\
&+ \sum_{l=2}^{n_L} \left( \frac{f_{lj} - h_{ljk}}{1 - g_{lk}} - \frac{f_{1j} - h_{1jk}}{1 - g_{1k}} \right) L_{ikl} \\
&+ 2 \cdot \frac{f_{1j} - h_{1jk}}{1 - g_{1k}}
\end{aligned} \tag{41}
$$

Combining this result with **Equation 33** yields **Equation 19**, thereby completing the proof.
